# The Pre-MIRACLE_2_ Score - Pre-hospital Risk Stratification of Resuscitated Out of Hospital Cardiac Arrest

**DOI:** 10.64898/2026.01.22.26344639

**Authors:** Muhamad Abd Razak, John Hodsoll, Prajith Jeyaprakash, Michael McGarvey, Garry Hamilton, Roman Roy, Evan Ansell, Sundeep Kalra, Peter Kordis, Antonio Cannata, Rupert Simpson, Uzma Sajjad, Nick. Curzen, Serena Rakar, Clare Appleby, Abdul Mozid, Satpal Arri, Krishnaraj Rathod, Piotr Pałczyński, Mariusz Sieminski, Julian Yeoh, Thomas W Johnson, Paul Rees, Thomas Keeble, Rafal Dworakowski, Rachael Fothergill, Marko Noc, Philip MacCarthy, Jonathan Byrne, Daniel Stahl, Nilesh Pareek

## Abstract

**Background:** Out-of-hospital cardiac arrest (OHCA) remains a global health burden where neurological injury sustained is a key predictor of mortality but there are challenges in early risk stratification. This study aims to derive the Pre-MIRACLE_2_ score, which excludes pH as a component from the MIRACLE_2_ score, as a means of stratifying neurological risk in a pre-hospital setting.

**Methods:** To validate the Pre-MIRACLE_2_ score, we used (i) the EUCAR Registry retrospectively analysed from 1 May 2012 to 31 December 2021, and (ii) the GLOBAL-MIRACLE Registry, a prospective cohort analysed from 1 January 2022 to 31 May 2023. The primary outcome was poor neurological outcome (defined as Cerebral Performance Category 3-5) at hospital discharge.

**Results:** From 1 May 2012 until 31 May 2023, 2149 patients were resuscitated from OHCA with sustained return of spontaneous circulation. After excluding patients who remained non-comatose following return of spontaneous circulation and those with incomplete scores, 1402 patients from EUCAR and 747 from GLOBAL-MIRACLE were included in the final analysis. The primary endpoint occurred in 54.4% of the study cohort. The performance of the Pre-MIRACLE_2_ score for the primary endpoint was excellent, with an area under the receiver operating curve (AUROC) of 0.85 (95% CI 0.83, 0.87). From the prospective validation cohort (GLOBAL-MIRACLE), the AUROC was 0.85 (95% CI 0.82-0.88) with a calibration slope of 1.11 (95% CI 0.95-1.29).

**Conclusion:** The Pre-MIRACLE_2_ score has the potential to be an effective and pragmatic risk stratification tool for prediction of poor neurological outcome in a pre-hospital environment or where the pH cannot be measured.

## Introduction

OHCA is a major public health burden and a leading cause of mortality globally.(1,2) Systematic pathways have been developed to expedite conveyance of patients resuscitated after OHCA direct to cardiac arrest centres (CAC) equipped with therapeutic interventions such as invasive coronary angiography and mechanical circulatory support (MCS), particularly for those in cardiogenic shock (CS).(3) While conceptually promising, randomised controlled trials (RCT) to date have failed to show benefit for direct conveyance to a CAC or for several interventions (4–7) and survival rates to discharge for OHCA from the community remain lower than 10% across many healthcare settings.(3,8) These neutral RCT results are potentially driven by a subgroup of this population with irreversible and significant neurological injury sustained prior to hospital arrival.(9) An inability to personalise risk stratification prior to conveyance might lead to transfer of inappropriate subsets of patients for specialist care, treatment futility unnecessary distress for patients/care-givers and inefficient distribution of healthcare resources such as critical care capacity.(10,11)

There is currently no established tool for personalised pre-hospital stratification of neurological risk in resuscitated OHCA.(12) Several risk tools such as Cardiac Arrest Hospital Prognosis (CAHP), Target Temperature Management (TTM), Out of Hospital Cardiac Arrest (OHCA) and NULL-PLEASE have been developed for use specifically on arrival to a centre (13–16) but these are often complex to administer and have been developed for application on critical care unit admission, a timepoint after conveyance decisions can be influenced.(14,17) Our group has previously developed and prospectively validated the MIRACLE_2_ score as a practical, point-based risk tool designed to be administered at the time of hospital admission for those with suspected cardiac aetiology.(18,19) With the exception of pH, all components of MIRACLE_2_ score can be readily and rapidly assessed pre-hospital by emergency medical services at the time of return of spontaneous circulation (ROSC). There is an unmet clinical need for an accurate, practical and well validated risk stratification tool in the pre-hospital setting of OHCA for stratification of neurological risk.

Accordingly, the purpose of this study was to determine whether a pragmatic modification of the MIRACLE_2_ score that excludes the pH variable, termed the “Pre-MIRACLE_2_ score”, can facilitate accurate pre-hospital prediction of neurological outcome for patients presenting with resuscitated OHCA of suspected cardiac aetiology, circumstances in which arterial blood gas data are typically unavailable.

## Methods

The study was performed according to the principles of the Declaration of Helsinki and was approved by local research ethics or governance committees at each centre for both EUCAR and GLOBAL-MIRACLE registries.

### Study Population

The derivation cohort represents the European Cardiac Arrest Registry (EUCAR) to retrospectively identify adult patients (aged >18 years) with ROSC maintained after OHCA in the community between 1 May 2012 and 31 December 2021. This registry has been established since 2012 and has complied with the standardised Utstein-style reporting template for OHCA.(20) Patients were included in this registry if they presented with ST-elevation Myocardial Infarction (STEMI), or if they lacked evidence for a non-cardiac cause of OHCA. Registry exclusion criteria included confirmed intra-cranial bleeding, neurological disability (CPC 3 or 4) prior to OHCA, presence of survival limiting illness, and a confirmed non-cardiac cause of OHCA or death prior to hospital arrival. Only patients with sustained ROSC and with successful conveyance to a centre were included in the registry. Any patients with intact conscious status [defined as a Glasgow Coma Score (GCS) of 15/15] on arrival were also excluded.

### Data Collection

Pre-hospital clinical data such as time of arrest, initial rhythm, bystander CPR and ROSC time were collected and medical records were accessed for biochemical and haemodynamic data upon hospital arrival. Baseline cardiac investigations such as electrocardiograms (ECG), echocardiograms and coronary angiograms were also collected where available from medical records.

### MIRACLE_2_ score derivation and validation and the Pre-MIRACLE_2_ score

The MIRACLE_2_ score is a practical risk tool developed in accordance with the TRIPOD (Transparent Reporting of a multivariable prediction model for Individual Prognosis Or Diagnosis) methodology.(18,21) The MIRACLE_2_ score consists of seven components (**Supplementary Table 1**) and was derived originally from 373 patients in the KOCAR registry with prospective validation in both the After-ROSC (22) and GLOBAL-MIRACLE registries (19) where it was shown to have at least equal if not superior discrimination to other proposed tools for predicting functional neurological outcome.

The Pre-MIRACLE_2_ score is a modification of the MIRACLE2 score which excludes the “L” (i.e. Low pH) parameter from the MIRACLE_2_ score to become a six variable and 9-point risk tool. As pH is the only biochemical marker requiring blood sampling, the remaining six parameters (1. Witnessed arrest, 2. Initial Rhythm, 3. Reactive Pupils, 4. Age, 5. Changing rhythm and 6. Epinephrine Dose) can be readily assessed by paramedics in the pre-hospital setting.

### Outcomes

The primary outcome measure for this study is poor neurological outcome, as defined by a CPC score of 3-5 on hospital discharge. A breakdown of the ordinal scale used to classify CPC is shown in **Supplementary Table 2**. The secondary outcome was circulatory-aetiology death, a combination of cardiac aetiology death and multi-organ dysfunction syndrome we have previously described (24).(23)

### External Validation

Validation was performed from the prospectively collected GLOBAL-MIRACLE registry which has the same study criteria as EUCAR. The GLOBAL-MIRACLE registry was a prospective multi-centre registry from 5 countries and 11 centres. It followed the same standardised Utstein-style reporting guidelines for OHCA with recruitment occurring from 1 January 2022 until 31 May 2023. The same primary endpoint of CPC score 3-5 at hospital discharge was assessed in this cohort.

### Statistical analysis and prediction modelling

Predictive models for the MIRACLE_2_ and Pre-MIRACLE_2_ scores were re-developed in the EUCAR cohort (n=1,402; original KOCAR cohort n = 373). External validation was performed in the GLOBAL-MIRACLE registry (n=747). Model performance was assessed in terms of discrimination (via AUC) and overall accuracy and fit (via Brier score and Nagelkerke’s R²).(24,25) Calibration was examined through loess-smoothed calibration plots and by estimating the calibration intercept and slope from linear regression models of observed versus predicted risk.

Internal validation for EUCAR was conducted using bootstrap resampling (200 replicates), with the rms package in R.(26) Bootstrapped samples were split into training and test sets to estimate optimism-corrected performance metrics. Comparative risk score performance between Pre-MIRACLE_2_ and MIRACLE_2_ was assessed using ROC curves and median paired differences in AUC (95% CI) in both derivation and validation cohorts. Additionally, patients were stratified into predefined risk categories based on the Pre-MIRACLE_2_ score (low: 0–2; moderate: 3–4; high: ≥5), and sensitivity, specificity, positive predictive value (PPV), and negative predictive value (NPV) were calculated for poor neurological outcome within each group.

Logistic regression explored the impact of low pH on poor neurological outcomes and circulatory-aetiology death, conditional on Pre-MIRACLE_2_ scores, with interaction terms. Logistic regression coefficients (log-odds) are reported, rather than odds ratios, to preserve interpretability in the presence of interaction terms and to reflect the additive structure of the model. We also assessed score changes from Pre-MIRACLE_2_ to MIRACLE_2_ due to an extra score fot the pH, examining reclassification rates across risk groups.

Due to missing predictor data, multiple imputation was conducted using Multiple Imputation (MI) by Chained Equations (MICE),(27) generating 50 imputed datasets. The imputation model derived from EUCAR was applied to GLOBAL-MIRACLE to ensure consistency. No outcome data were imputed. Model optimism metrics for internal validation were averaged across 50 imputations and the across-imputation standard deviation as an index of the stability of these metrics. 95% confidence intervals (CI) for the model performance metrics were generated by combining 500 bootstrap samples for each imputation dataset and calculating 2.5% and 97.5% percentiles.(28) For the exploratory modelling of Pre-MIRACLE_2_ score by Low pH interactions, statistical significance of the parameters was set at p < 0.05 and parameters were presented with 95% CIs. All analyses were undertaken using R version 4.5.0. Full statistical details are shown in the **supplementary material**.

## Results

From 1st May 2012 to 31st May 2023, 2339 patients resuscitated from OHCA were included in EUCAR and the GLOBAL-MIRACLE registries. After excluding patients who were non-comatose (GCS 15/15) (n=190) following ROSC, 2149 patients (1402 in the derivation cohort and 747 in the validation cohort) were included in the final analysis (**Figure 1**). Of these, n= 249 (12%) of patients had incomplete pre-hospital variables to calculate the Pre-MIRACLE_2_ score. Eight patients (0.57%) had missing outcomes and so were removed from further analysis beyond descriptive statistics.

**Figure 1.**
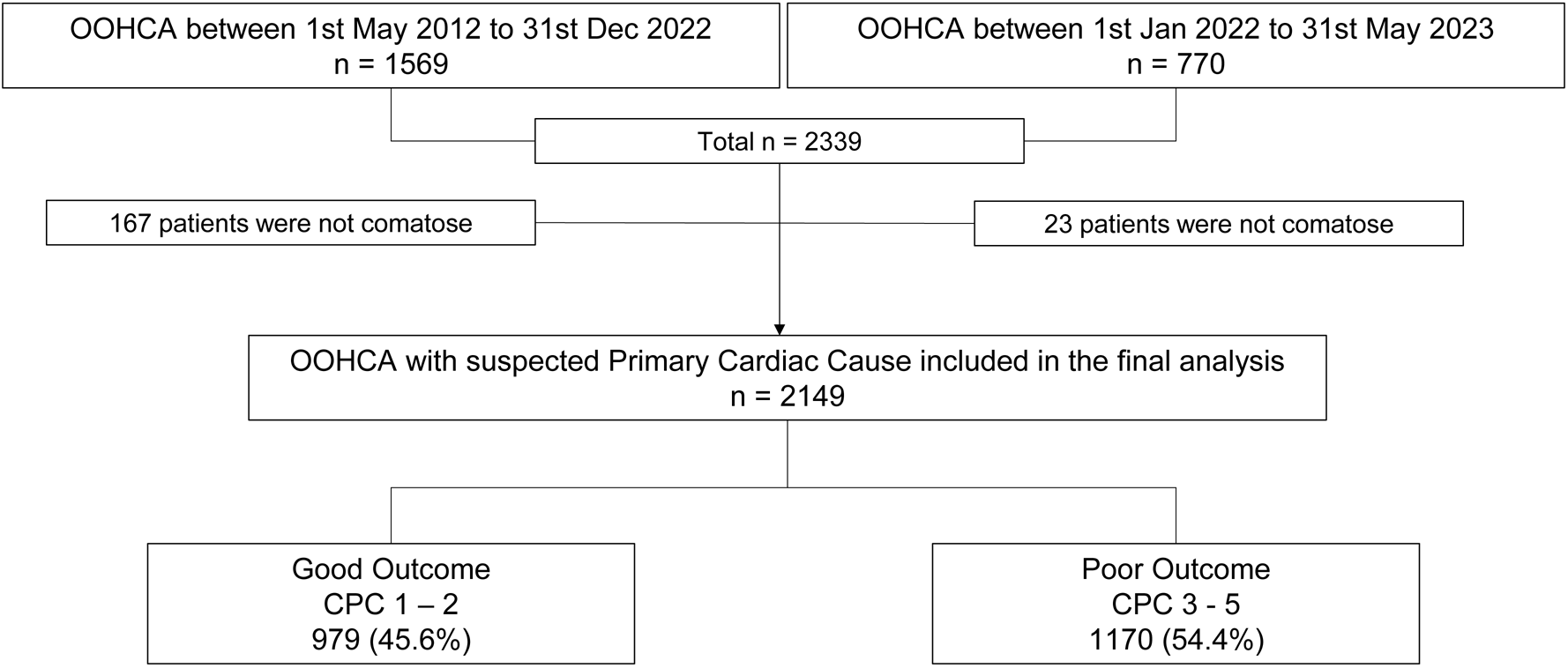
Study Flow chart.

From the derivation study, the median age was 63 years (IQR 53-73) and the majority of patients (78%) were male. One thousand and fifteen patients (76%) had an initial shockable rhythm, 79% had witnessed OHCA and 75% of patients had bystander CPR. The median zero and low-flow times were 0 minutes (IQR 0-5) and 20 minutes (IQR 12-31) respectively. On arrival to a centre, seven hundred and thirty-nine patients had ST-segment elevation (STEMI) or left bundle branch block (LBBB) on post-ROSC 12-lead ECG and 358 patients had immediate coronary angiography (CAG) on arrival to the cardiac arrest centre with 45.8% having percutaneous coronary intervention (PCI) (**Table 1**). The primary outcome measure of poor neurological outcome (CPC 3-5) at hospital discharge occurred in 785 patients (56.3%) (**Table 1)**

**Table 1.**
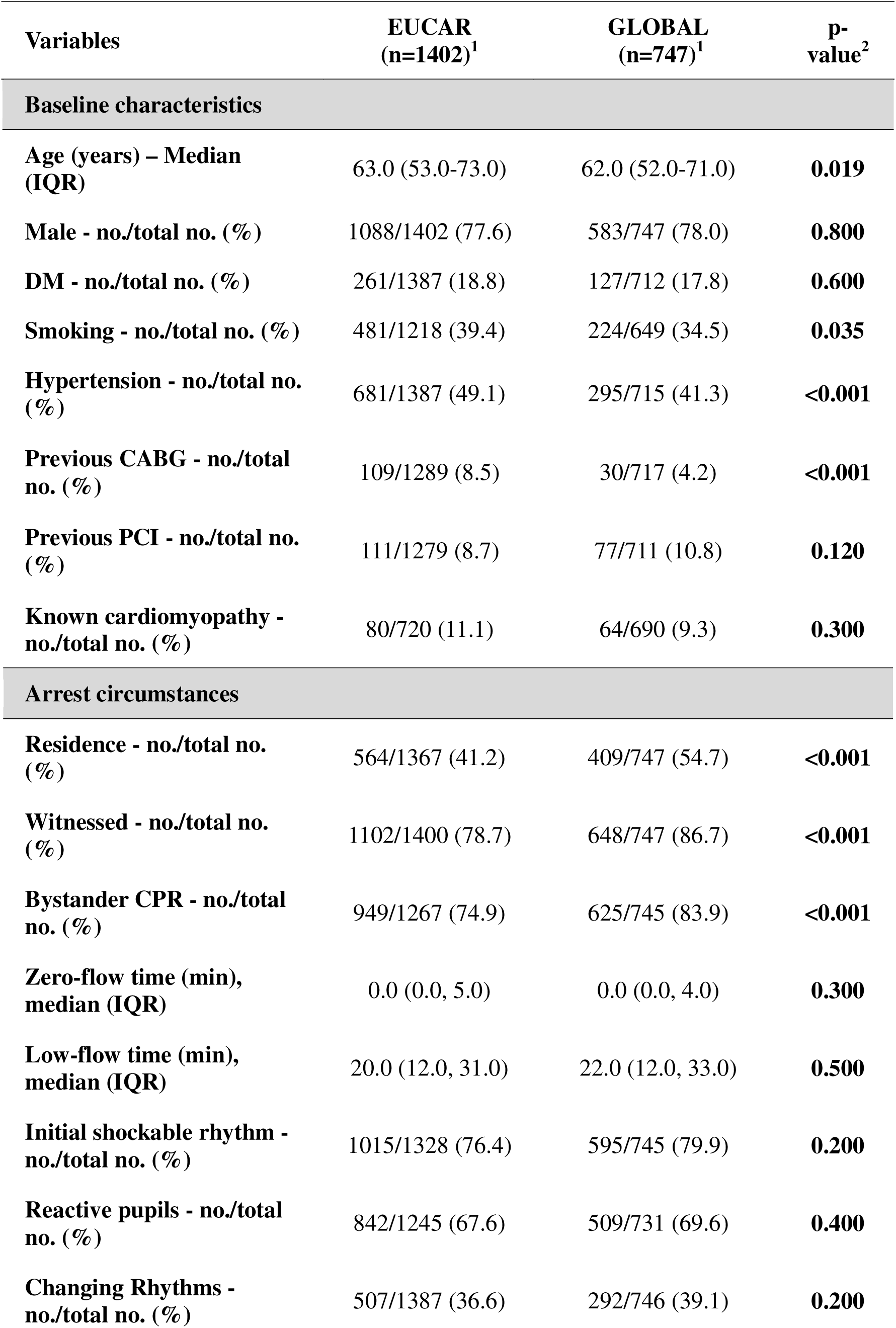
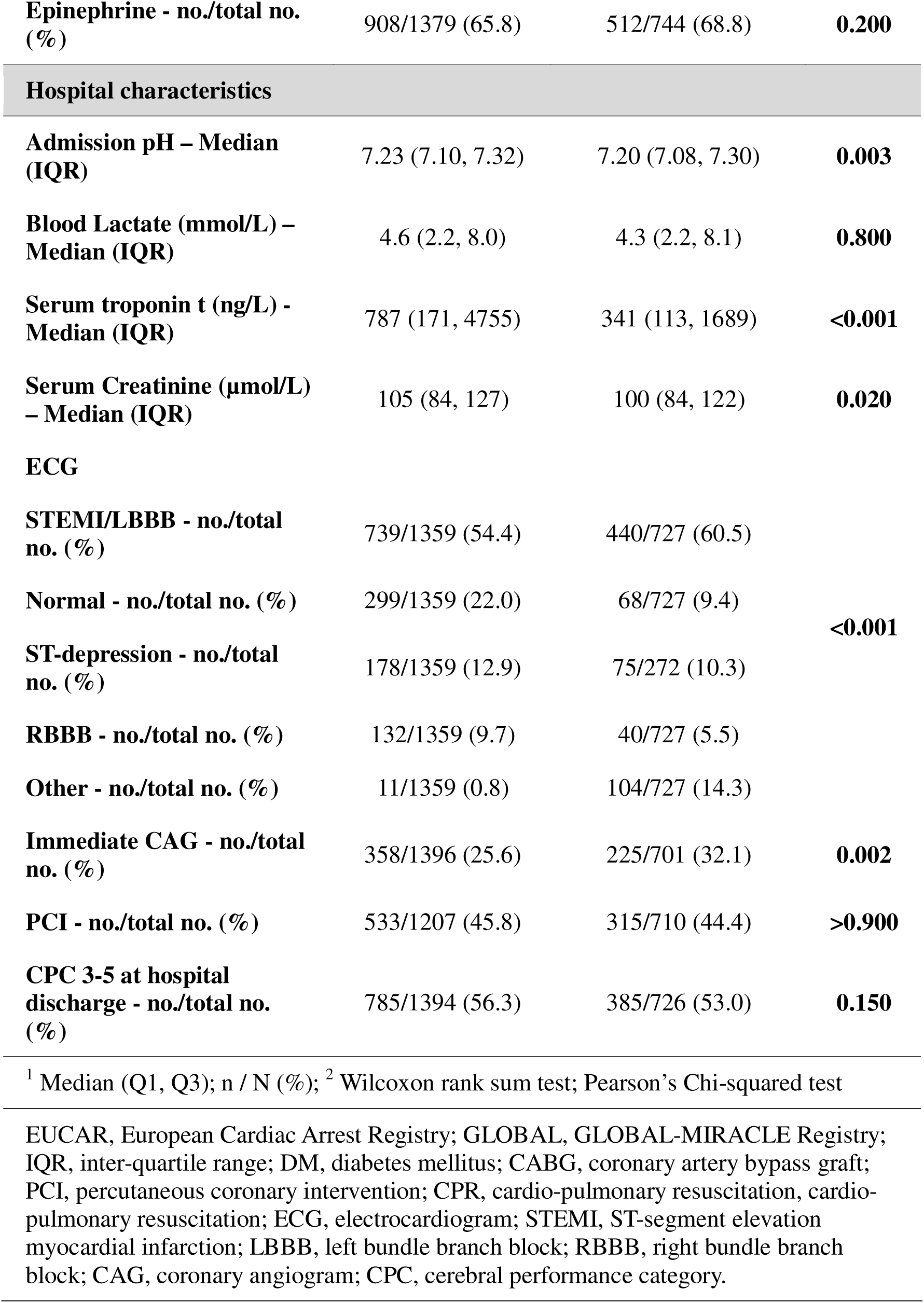
Descriptive statistics for key demographic and clinical variables for EUCAR and GLOBAL datasets.

### Internal Performance of the Pre-MIRACLE_2_ score in predicting poor neurological outcome

In EUCAR, internal validation showed negligible optimism (<0.002 for all metrics) across imputed datasets (**Supplementary Table 3**) and so uncorrected performance statistics are presented. The Pre-MIRACLE_2_ score had an AUC of 0.85 (95% CI 0.83-0.87) with a R2 of 0.46 (95% CI 0.42-0.52) and Brier score of 0.15 (95% CI 0.14-0.16). As with the MIRACLE_2_ score, there was a step-wise increase in the primary endpoint of poor neurological outcome as the Pre-MIRACLE_2_ score increased (p<0.0001) (**Central Illustration, Figure 2 and Table 2**). Calibration was excellent both in terms of model estimation and visually (**Table 2)**. Pre-MIRACLE_2_ appeared to slightly underestimate the risk of poor outcome at higher Pre-MIRACLE_2_ scores compared with the MIRACLE_2_ score **Figure 2**.

**Figure 2.**
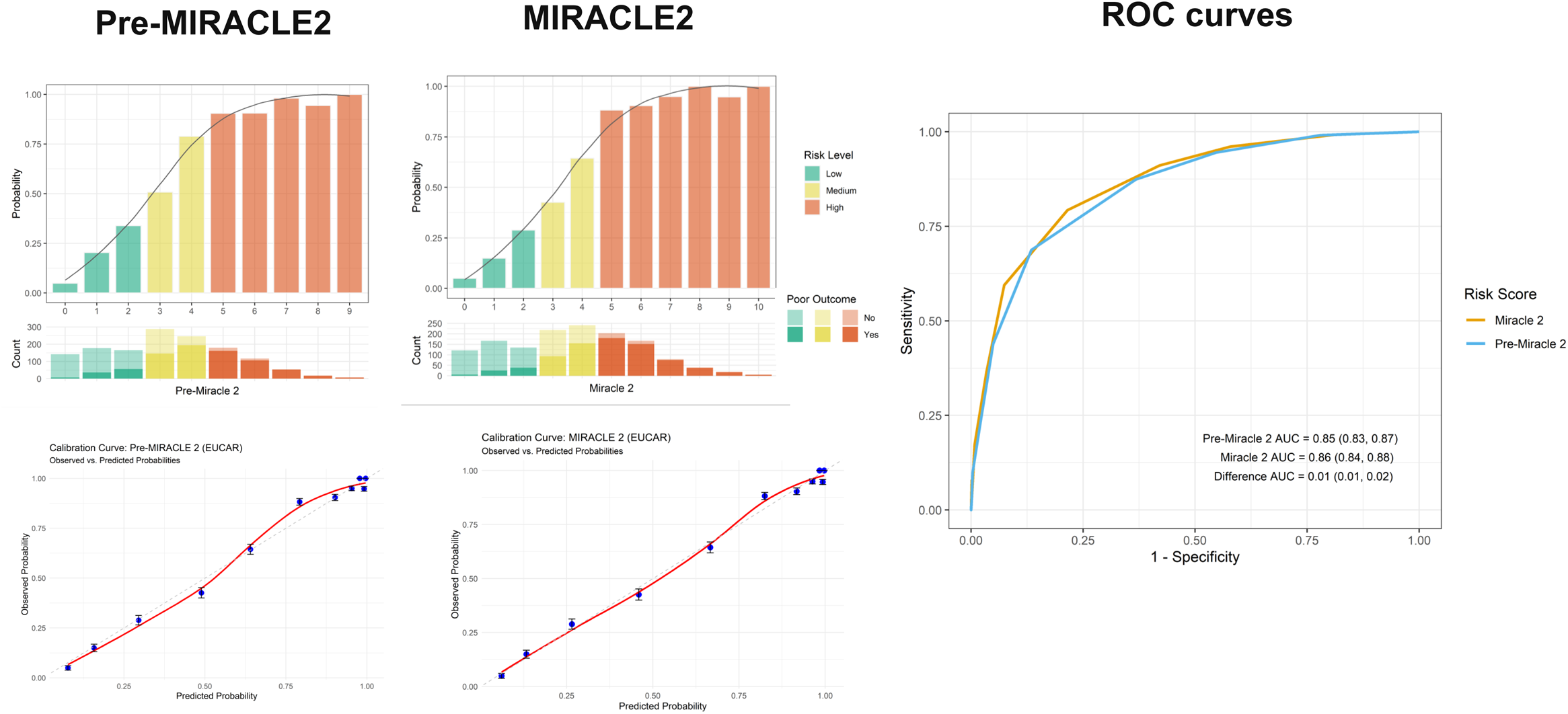
Performance of Pre-MIRACLE_2_ and MIRACLE_2_ in the EUCAR cohort. Observed outcome probabilities across score strata are shown for Pre-MIRACLE_2_ and MIRACLE_2_, with corresponding score distributions. Receiver operating characteristic curves demonstrate comparable discrimination between scores, with similar areas under the curve.

**Table 2.**
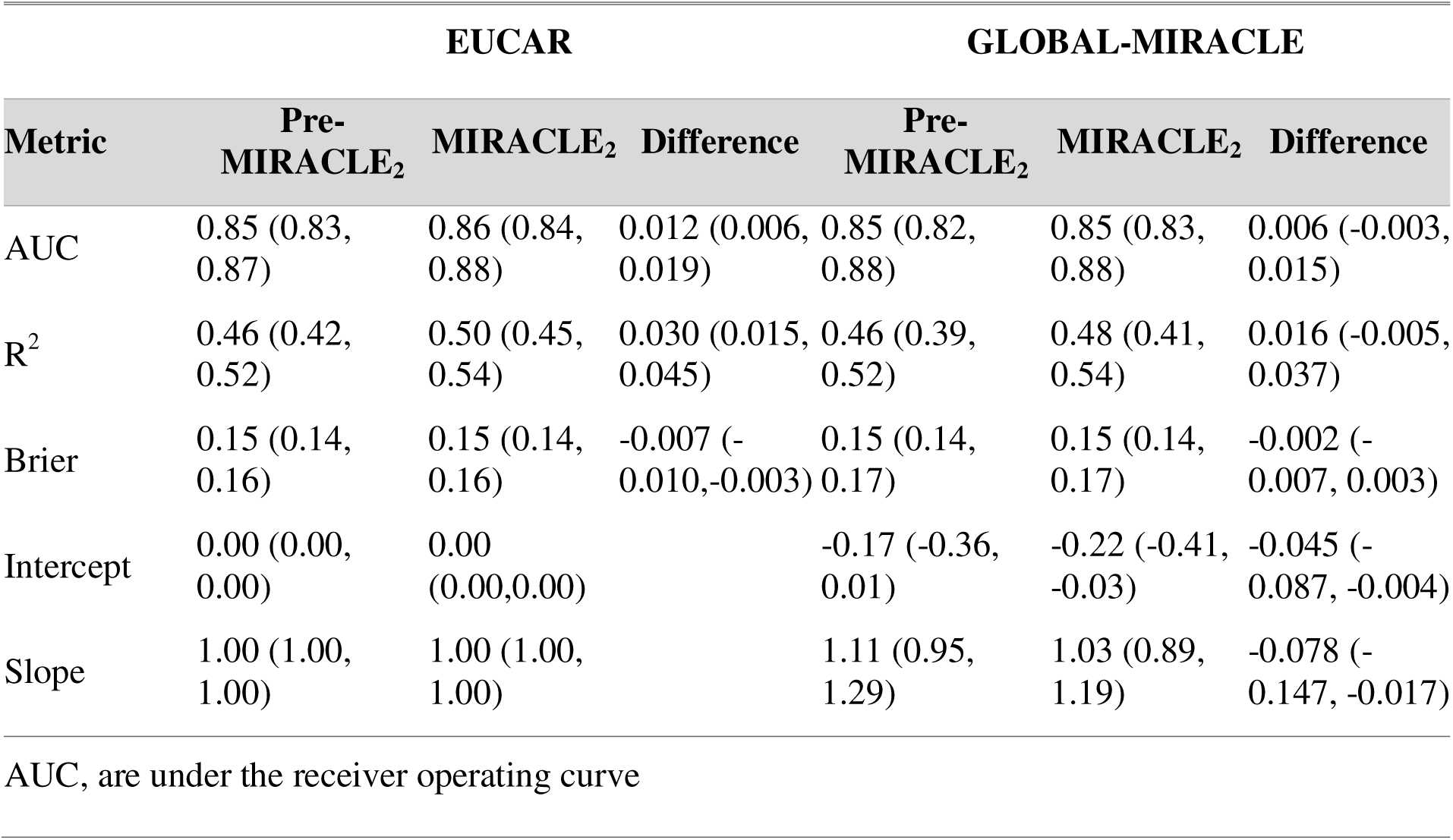
Performance metrics by risk score and cohort (EUCAR and GLOBAL), estimated AUC, R^2^, Brier Score and Calibration Slope and Intercept across multiply imputed datasets (m = 50) with 95% bootstrapped confidence intervals (B = 500 per imputation)

### External Validation Cohort

Seven hundred and forty-seven patients from the GLOBAL-MIRACLE registry cohort were included in this analysis with baseline demographics and cardiac arrest circumstances listed in **Table 1**. Patients in the GLOBAL-MIRACLE registry were similar to EUCAR other than lower rates of hypertension, prior CABG, but higher rates of arrests occurring in a residential location, witnessed or receiving bystander CPR. In the GLOBAL-MIRACLE cohort, the AUC for the Pre-MIRACLE_2_ score was 0.85 (95% CI 0.82-0.88) with a calibration slope of 1.11 (95% CI 0.95-1.29) (**Figure 3**). Again, calibration was excellent (**Table 2 and Figure 3**). Visually below observed probabilities of approximately 0.6, Pre-MIRACLE_2_ over-estimates the risk of a poor outcome.

**Figure 3.**
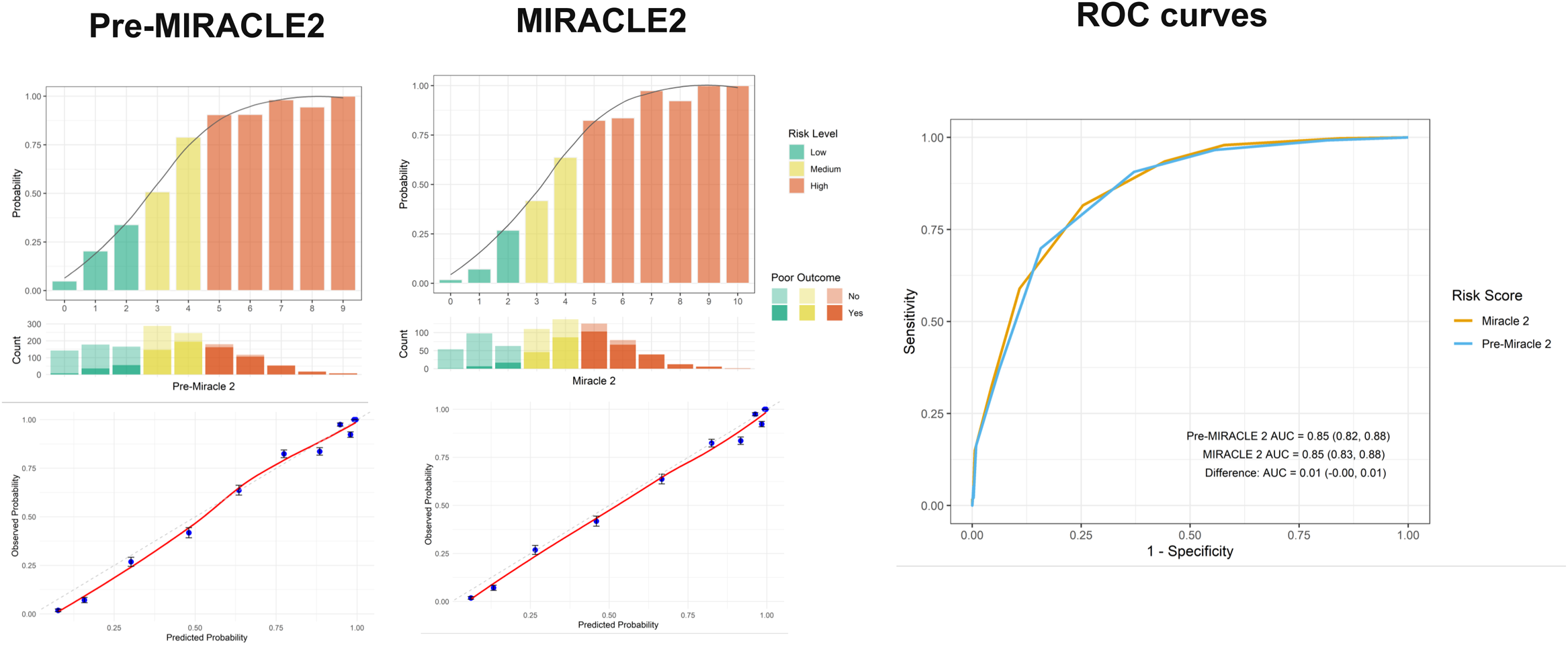
Comparison of Pre-MIRACLE_2_ and MIRACLE_2_ performance in GLOBAL-MIRACLE. Observed outcome probabilities across score strata are shown for Pre-MIRACLE_2_ and MIRACLE_2_, with corresponding score distributions. ROC curves (right panel) show similar discriminative performance for Pre-MIRACLE_2_ and MIRACLE_2_ (AUC 0.85 for both; difference 0.01, 95% CI −0.00 to 0.01).

### Discrimination performance of the Pre-MIRACLE_2_ score by risk groups

Three groups were identified Pre-MIRACLE_2_ score (low – Pre-MIRACLE_2_ score 0-2; intermediate – 3-4; high – ≥5). In patients with complete data in EUCAR, 501/1285 (39%) were in the low-risk group, 463/1585 (36%) in the intermediate and 327 (25.4%) in the high-risk group. The primary endpoint occurred in 76/499 (15%) of the low-risk group, 291/463 (63%) in the intermediate group and 297/325 (91%) of the high-risk group. Patients at low risk (Pre-MIRACLE_2_ ≤ 2) had a negative predictive value of 0.80 (0.76, 0.83) in EUCAR and 0.86 (0.81, 0.90) in GLOBAL-MIRACLE. Patients with high risk (Pre-MIRACLE_2_ ≥ 5) had a high risk of poor neurological outcome with a positive predictive value of 0.92 (0.89, 0.95) in EUCAR and 0.87 (0.82, 0.92) in GLOBAL-MIRACLE. A Pre-MIRACLE_2_ score of ≥7 had a positive predictive value of 0.92 (0.89, 0.95) and 0.95 (0.84, 1.00) in the EUCAR and GLOBAL-MIRACLE registries respectively (**Table 3**, **Figures 2 and 3 and Supplementary Figure 1**.

**Table 3.**
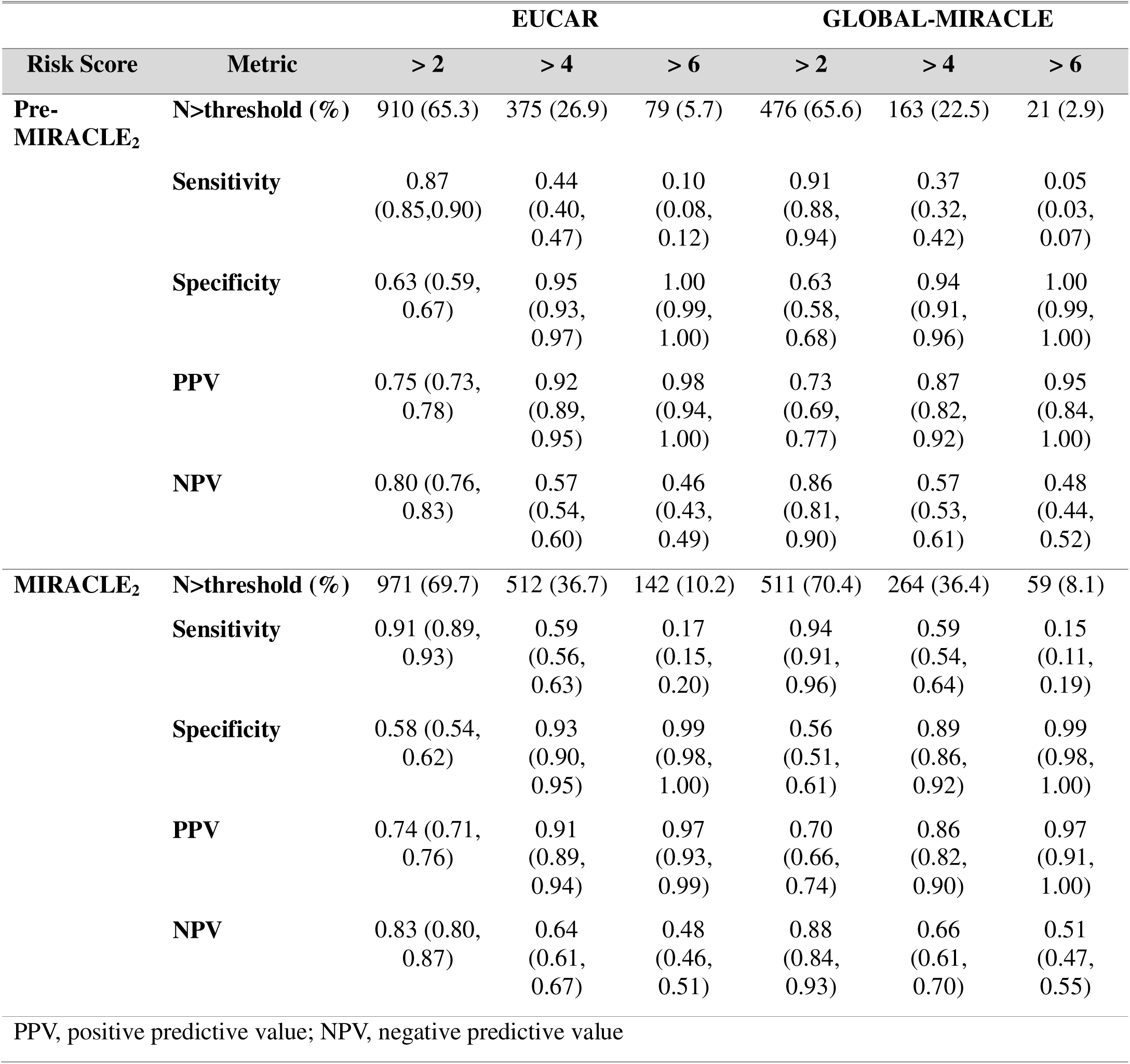
Estimates of sensitivity, specificity, positive predictive values and negative predictive values by risk category threshold for Pre-MIRACLE_2_ and MIRACLE_2_ scores by cohort.

### Comparison of Model fit of Pre-MIRACLE_2_ and MIRACLE_2_ scores

The MIRACLE_2_ score shows slightly superior discrimination than the Pre-MIRACLE_2_ score in EUCAR [AUC 0.86 (0.84, 0.88), difference 0.012 (0.006, 0.019)] but equivalent performance in GLOBAL-MIRACLE [AUC 0.85 (0.83, 0.88), difference 0.006 (−0.003, 0.015)]. For EUCAR there was equivalent calibration determined by model of predicted versus observed outcomes, both intercepts = 0 and slope = 1. In the validation GLOBAL-MIRACLE registry, MIRACLE_2_ also had superior calibration than Pre-MIRACLE_2_ in slope [1.03 (0.89, 1.19), difference −0.08 (−0.15, −0.02)] and intercept [-0.22 (−0.41, −0.03), difference −0.045 (−0.087, −0.004)]. Finally, incorporation of a low pH into the MIRACLE_2_ score also improves model fit with model comparison using the Likelihood Ratio Test for MI (D3). The test comparing the model with low pH as a predictor compared to the nested model without yielded a statistically significant result, D3 (1, 1.13 x 10^8^) = 35.52, p < 0.001.

### Change in Pre-MIRACLE_2_ score to MIRACLE_2_ score and effect on outcome

A total of 614 patients (44.0%) in the EUCAR analysis set accrued an additional point (due to low pH) between the Pre-MIRACLE_2_ and MIRACLE_2_ scores. As the Pre-MIRACLE_2_ score increased, the probability of a low pH increased. Dynamic change in the score was more prevalent as risk group increased: this occurred in 113 (23.3%) in the low-risk group, 379 (53.2%) of the intermediate group and 122 (61.9%) of the high-risk group. Within the low risk group, inclusion of low pH led to 61 (12.4%) switching to intermediate risk and 131 (17.8%) of the intermediate group switching to the high risk group (**Supplementary Figure 2**).

#### Effect on poor neurological outcome

In a multivariable logistic regression model for poor neurological outcome on hospital discharge, the Pre-MIRACLE_2_ score had a log odds ratio (OR) of 0.81 (95% CI 0.70-0.93) and a low pH had a log odds ratio of 0.71 (95% CI 0.15-1.5) (p 0.022). The interaction of the 2 variables was not significant [0.07 (95% CI −0.14 −0.27 (p 0.5)] indicating that the effect of an additional point of a low pH on arrival was independent of the Pre-MIRACLE_2_ score on the log-odds scale for the primary endpoint of poor outcome (**Figure 4A**).

**Figure 4.**
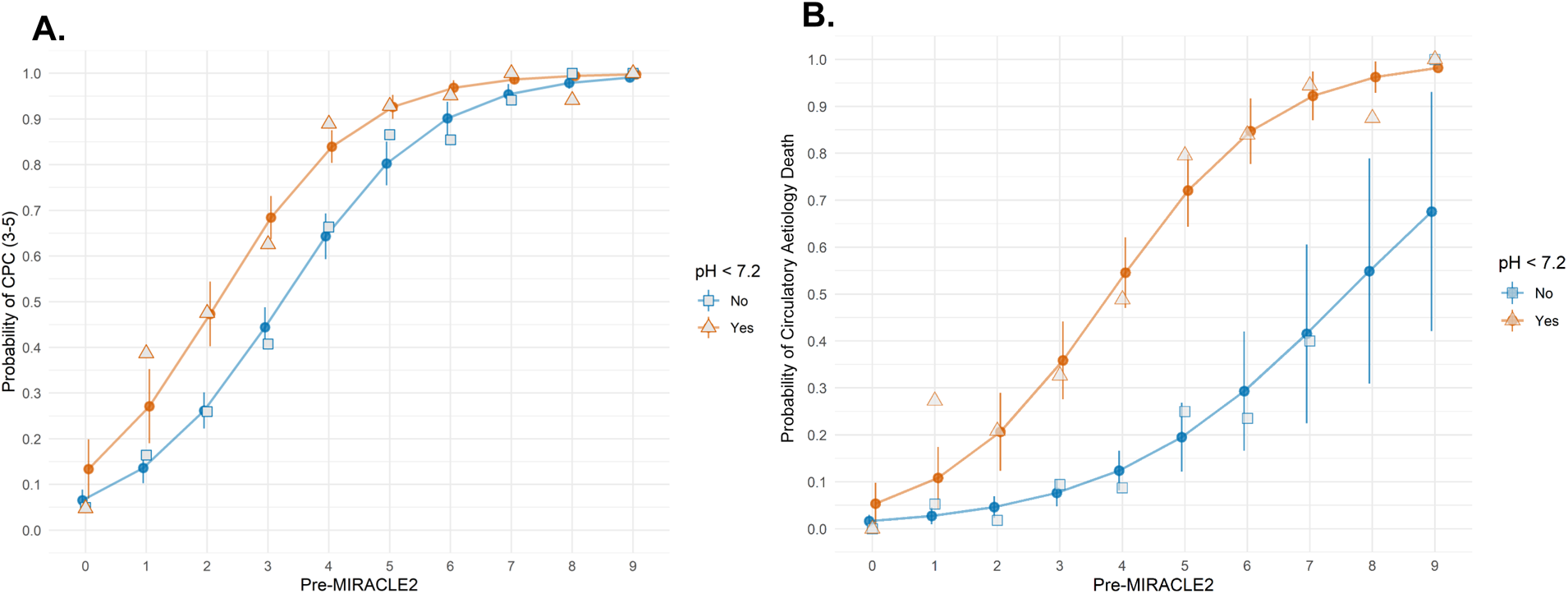
Association Between Pre-MIRACLE_2_ Score and Predicted Probability of Poor Neurological Outcome and Circulatory Aetiology Death, Stratified by Arterial pH <7.2. Predicted probability of poor neurological outcome (CPC 3–5) (Panel A) and Circulatory Aetiology Death (Panel B) according to Pre-MIRACLE_2_ score, stratified by arterial pH <7.2. Blue squares/triangles: pH ≥7.2 (No); orange triangles: pH <7.2 (Yes).

#### Effect on circulatory aetiology death

For patients with complete data for the secondary endpoint of circulatory aetiology death (n = 641), a low pH had a log OR^1^ of 1.2 (95% CI 0.00, 2.5) (p 0.051). An interaction analysis with Pre-MIRACLE_2_ was also non-significant [0.23 (95% CI −0.07, 0.53) (p 0.14]), indicating again an independent association of gaining an additional point for a low pH on circulatory aetiology death (**Figure 4B**).

## Discussion

In this study, we have developed the Pre-MIRACLE_2_ score through modification of the MIRACLE_2_ score in a retrospective cohort of resuscitated OHCA (EUCAR) with subsequent validation in the prospective GLOBAL-MIRACLE registry. The Pre-MIRACLE_2_ score was almost as effective at neurological risk stratification as the hospital MIRACLE_2_ score, but with ease of calculation at the time of ROSC. When applied in a pre-hospital setting prior to conveyance, the Pre-MIRACLE_2_ score may be an effective and practical risk stratification tool for neurological outcome in patients with resuscitated OHCA with suspected cardiac aetiology.

The importance of early risk stratification for resuscitated OHCA is increasingly recognised. Recent American Heart Association (AHA) guidelines emphasised that risk tools might have an important role to play in future practice but should be reliable, easy to use and “applicable at the patient bedside”.(29) In a joint statement from the neurocritical care society and the AHA, the importance of an early risk stratification tools to “facilitate triage, guide family conversations and resource use” in the setting of OHCA is highlighted.(11) Several national and international societies have also emphasised the need for system based quality improvement in OHCA care.(30,31) While the ARREST trial did not show any benefit for expedited conveyance of those without STEMI to cardiac arrest centres,(32) OHCA is highly heterogeneous and patients with irreversible neurological injury may have negated any potential positive impact in a lower risk group. Pre-hospital stratification of underlying neurological risk combined with personalised pathways of care may identify subsets of patients with most to gain from expedited conveyance to specialist centres. Conversely, a robust risk tool that identifies genuine clinical futility could save both healthcare resource and emotional stress for family members.

Several risk tools, such as the CAHP, OHCA and TTM scores,(13–15) have been developed and validated for implementation after admission to a centre, usually on arrival to the intensive care unit (ICU). However, these complex algorithms usually incorporate biochemical data and other detailed information such as duration of downtime that are not readily available in a pre-hospital setting. More complex tools employing machine learning algorithms such as the SCARS-1 and by Tatesihi et al. have been developed with good performance.(33,34) However, these are complex algorithms, with multiple variables, designed for use in both those with and without ROSC, and so unlikely to gain widespread clinical usage in an emergency setting. By contrast, the Pre-MIRACLE_2_ score can be feasibly performed by emergency medical services prior to hospital transfer without the need for biochemical or chronological parameters. Early presentation of the Pre-MIRACLE_2_ score has been performed in small studies in a pre-hospital setting from both a helicopter emergency medical service cohort or in those ultimately conveyed to intensive care unit have also shown promising performance, but without formal prediction modelling.(35,36)

In this study, the Pre-MIRACLE_2_ score, in both the derivation and prospective validation cohorts had an AUC of 0.85 with satisfactory calibration. Stratification of Pre-MIRACLE_2_ into a low-risk group had a NPV of 0.86, indicating a high chance of good neurological whereas those in the high-risk group (≥5) had a PPV of poor neurological outcome of 0.87. Those at highest risk (Pre-MIRACLE_2_ ≥7) had a PPV of >95% indicating a near futile cohort of patients identified at the time of ROSC.

The Pre-MIRACLE_2_ score has similar prediction of poor neurological outcome as the MIRACLE_2_ score, though the MIRACLE_2_ score has superior discrimination in EUCAR and also superior calibration and model fit, particularly seen in the prospective GLOBAL-MIRACLE validation cohort. This raises the question of the importance of measuring the metabolic status of the patient, by collecting pH, on arrival to a centre and any added value of a low pH for classification of outcome. It is well established that a lower pH arising from a combination of metabolic and respiratory acidosis portends a poor prognosis, indicating several deleterious processes including poor ventilatory status and tissue hypoperfusion, while itself exacerbating irreversible cellular damage.(37–39) It is possible that the Pre-MIRACLE_2_ score incorporates a collection of variables which accurately reflect, as a surrogate, the metabolic status of the patient. However, the degree of acidaemia, and particularly lactataemia, also reflects the cardiogenic shock state of the patient on arrival to a centre and we propose that arterial blood gas analysis should remain an important initial hospital investigation in this patient group. Determination of arterial blood levels can be increasingly performed in the pre-hospital setting but the discrimination and clinical relevance of pH in this setting remains less clear. Our group is currently undertaking the RAPID-MIRACLE study which is evaluating the use of point of care pre-hospital pH and calculation of the MIRACLE_2_ score and is due to report its findings in due course (REC 22/EE/0001).

In this study, we found that a proportion of patients are re-classified from the initial Pre-MIRACLE_2_ score to the MIRACLE_2_ score based on a low pH on arrival to a centre. Patients in the highest risk group (Pre-MIRACLE_2_ ≥5) were most likely to be reclassified to a higher risk score on arrival, indicating that they have already sustained a more severe systemic tissue injury with organ hypoperfusion prior to arrival to a centre. Irrespective of the underlying score, gaining a point by transitioning from Pre-MIRACLE_2_ to MIRACLE_2_ by virtue of low pH was associated with poor neurological outcome and circulatory aetiology death. If used as part of the handover process on arrival to a centre, this might be used as a guide to highlight patients at higher risk of haemodynamic instability, but this notion requires further validation.

The Pre-MIRACLE_2_ score has several important implications for this condition, potentially on a global scale. With prospective validation, the Pre-MIRACLE_2_ score has the potential to guide personalised early stratification for expedited conveyance to cardiac arrest centres. While the ARREST trial failed to show benefit for this strategy in those without STEMI,(32) this trial population had a relatively rate of non-shockable rhythms and high inherent neurological risk. A recent observational registry of implementation of the BCIS protocol for expedited conveyance of those with either STEMI or a shockable rhythm showed an improvement in outcome for those with an initial shockable rhythm and a low-intermediate MIRACLE_2_ score (≤5).(3,40) Prospective validation of this approach in RCTs across differing healthcare systems and population densities is now required to confirm this preliminary finding. The Pre-MIRACLE_2_ score may also play an important role in identifying appropriate OHCA patients at low risk of neurological injury for direct referral to Level 1 cardiogenic shock centres to avoid secondary transfers and to maximise critical care capacity. Importantly, the Pre-MIRACLE_2_ score may also be of particular use in guiding individualised pre-hospital interventions, such that those at low risk might be prioritised for advanced critical support and more intensive cardiovascular therapies while those at high risk might be considered for novel investigational neuro-protective therapeutic approaches. Finally, identifying patients at low risk of neurological injury with the Pre-MIRACLE_2_ score for expedited conveyance to specialist centres may have potential for wider spread global application in large geographical areas or in low income countries, where the infrastructure for specialist care remains costly and resources, such as advanced MCS, must be prioritised.

## Limitations

Although the derivation of the Pre-MIRACLE_2_ score was from a large multi-centre registry, this was retrospective in nature and included suspected cardiac causes of arrest with a GCS<15/15, which is reflected in the high rates of initial shockable rhythms. Patients from both the EUCAR and GLOBAL-MIRACLE registries were also conveyed and treated in specialist cardiac arrest centres. Importantly, for this reason, the score should be applied only in those with comatose OHCA with sustained ROSC and with reasonable and realistic consideration for conveyance to a centre. The Pre-MIRACLE_2_ score may not be as reproducible in non-cardiac OHCA with higher rates of non-shockable rhythms, or in centres where advanced MCS support is not available and requires validation in these cohorts. Nevertheless, the findings from this study are highly relevant to this population as a practical pre-hospital risk stratification tool. Finally, it should also be emphasised, in with recent AHA guidelines, that Pre-MIRACLE_2_ should serve only as an early risk stratification tool and not a means of formal neuro-prognostication on an individual level. Guidelines continue to indicate that this should occur at the 72 hour period after the event prior to withdrawal of life sustaining therapy.(41) Inclusion in these registries mandated that patients survive until arrival to hospital, whereas this risk tool would be applied in the community to facilitate decision-making regarding conveyance. Hence, prospective validation on scene and in randomised controlled trials is required before the Pre-MIRACLE_2_ score is considered for guideline incorporation of conveyance pathways in OHCA.

## Conclusion

The Pre-MIRACLE_2_ score has the potential to be an effective pragmatic and personalised risk stratification tool for prediction of poor neurological outcome in a pre-hospital environment or where the pH cannot be measured.

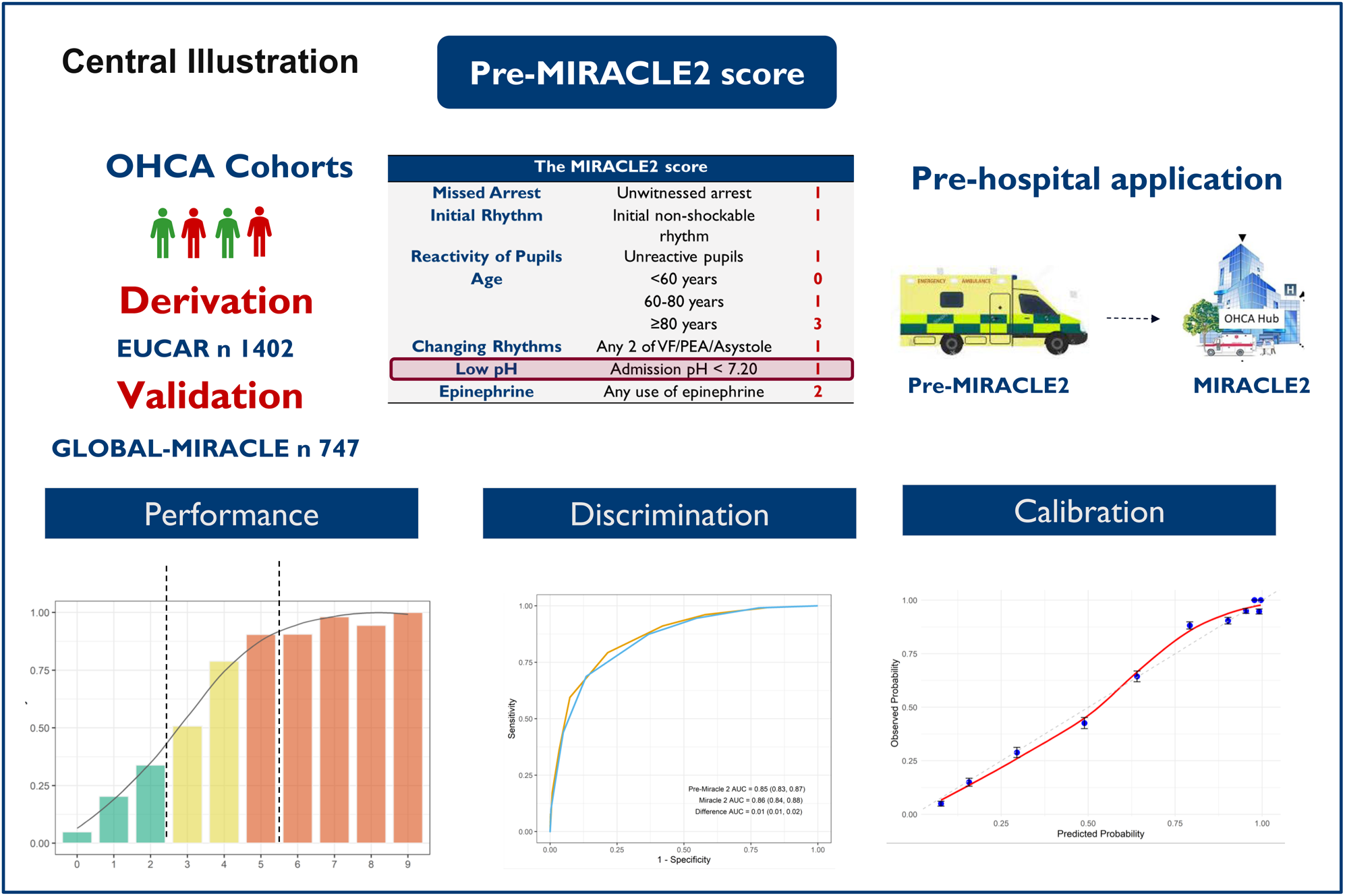
**Central Illustration. The Pre-MIRACLE2 score.**

## Abbreviations

CAC: Cardiac attack centre
CPC: Cerebral performance category
CPR: Cardiopulmonary resuscitation
MCS: Mechanical circulatory support
OHCA: Out-of-hospital cardiac arrest
PCI: Percutaneous coronary intervention
RCT: Randomised controlled trial
ROSC: Return of spontaneous circulation
SCAI: Society for cardiovascular angiography and interventions
STEMI: ST-elevation myocardial infarction

## Data Availability

All data produced in the present study are available upon reasonable request to the authors.

