## Supplementary Files for "The Pre-MIRACLE_2_ Score - Pre-hospital Risk Stratification of Resuscitated Out of Hospital Cardiac Arrest"

**Statistical Analysis Details**

**Dataset and Imputation**

The EUCAR dataset (n=1402), an update of the KOCAR dataset (n=373), was used for model development. The GLOBAL dataset, with no overlapping patients, served for external validation. Missing predictor values were handled by generating 50 imputed datasets using Multiple Imputation by Chained Equations (MICE, Van Buuren et al., 2011). The EUCAR imputation model was applied to the GLOBAL dataset to ensure consistency. No imputation was performed for missing outcome data.

**Model Development and Performance Metrics**

Predictive models for MIRACLE_2_ and Pre-MIRACLE_2_ risk scores were re-developed using the EUCAR dataset. Discrimination was evaluated using Area Under the Curve (AUC), Brier scores, and R². Optimism was assessed via bootstrapping (rms package, Harrell, 2009), splitting bootstrap samples of imputed datasets into training and test sets to estimate performance differences.

**External Validation**

In the GLOBAL dataset, model performance was assessed using AUC, Brier scores, R², and calibration. Calibration was evaluated visually with loess-smoothed fitted lines of predicted versus actual probabilities and statistically via the intercept and slope of a linear regression. Performance metrics were pooled across 50 imputations and 500 bootstrap re-samples (MI-BOOT, Schomaker et al., 2018), yielding 25,000 samples for percentile confidence intervals.

**Comparative Analysis**

Performance of MIRACLE_2_ and Pre-MIRACLE_2_ was compared using ROC curves and median differences (with 95% CI) in AUC, Brier scores, and R² for EUCAR and GLOBAL datasets. Calibration differences in GLOBAL were assessed visually and via bootstrap samples. Risk groups (low: 0-2, medium: 3-4, high: ≥5) were used to compute sensitivity, specificity, positive predictive value (PPV), and negative predictive value (NPV) at category thresholds for both scores.

**Logistic Regression Analysis**

Logistic regression models included an interaction term between low pH and Pre-MIRACLE_2_ score to assess the impact of low pH on poor neurological outcomes (Cerebral Performance Category 3–5) and circulatory death. Predicted probabilities were compared between groups with and without low pH, conditional on Pre-MIRACLE2 scores.

**Software**

All analyses were performed using R version 4.5.0, with packages including rms (Harrell, 2009) for bootstrapping and MICE (Van Buuren et al., 2011) for imputation.

| Supplementary Table 1: MIRACLE_2_ score components (1) | | | |
| --- | --- | --- | --- |
|  | **Variable** | **Definition** | **Points** |
| M | **M**issed | Unwitnessed Arrest | 1 |
| I | **I**nitial Rhythm | Non-shockable Rhythm | 1 |
| R | **R**eactive Pupils | No Pupil reactivity on ROSC | 1 |
| A_2_ | **A**ge | >60  >80 | 1  3 |
| C | **C**hanging Rhythms | Any 2 of VF/PEA/Asystole | 1 |
| L | **L**ow pH | pH <7.20 | 1 |
| E | **E**pinephrine | Any Adrenaline Dose | 2 |
|  |  | MIRA_2_CLE_2_ Score | 10 |

| **Supplementary Table 2. Cerebral Performance Category (CPC) ordinal scale for neurological impairment (2)** | |
| --- | --- |
| **CPC 1** | Good cerebral performance (normal life). Conscious; alert, able to work. May have minor neurologic or psychologic deficit (mild dysphagia, non-incapacitating hemiparesis, or minor cranial nerve abnormalities). |
| **CPC 2** | Moderate cerebral disability (disabled but independent). Conscious; sufficient cerebral function for independent activities of daily life. Able to work in sheltered environment. May have hemiplegia, seizures, ataxia, dysarthria, dysphasia, or permanent memory or mental changes. |
| **CPC 3** | Severe cerebral disability (conscious but disabled and dependent). Conscious; dependent on others for daily support because of impaired brain function. Limited cognition. Ranges from ambulatory state to severe dementia or paralysis. |
| **CPC 4** | Coma/vegetative state (unconscious). Unconscious, unaware of surroundings, no cognition. No verbal or psychologic interaction with environment. |
| **CPC 5** | Brain death. Certified brain dead or dead by traditional criteria. |

| **Supplementary Table 3. Bootstrap estimates of out-of-sample optimism** | | | |
| --- | --- | --- | --- |
| Metric | Apparent^1^ | Optimism^1^ | Corrected^1^ |
| Pre-Miracle | | | |
| AUC | 0.850 (0.000) | 0.0000 (0.0007) | 0.850 (0.001) |
| Brier Score | 0.154 (0.000) | -0.0004 (0.0004) | 0.154 (0.000) |
| R² | 0.465 (0.000) | 0.0004 (0.0018) | 0.465 (0.002) |
| Miracle 2 | | | |
| AUC | 0.863 (0.000) | -0.0001 (0.0008) | 0.863 (0.001) |
| Brier Score | 0.147 (0.000) | -0.0004 (0.0005) | 0.147 (0.000) |
| R² | 0.495 (0.000) | 0.0001 (0.0020) | 0.495 (0.002) |
| ^1^Mean (SD) across 50 imputed datasets | | | |

**Supplementary Figure 1. Performance metrics of the Pre-MIRACLE2 and MIRACLE2 risk scores for predicting poor neurological outcome after OHCA.** Panels display positive predictive value (PPV), negative predictive value (NPV), sensitivity, and specificity across risk score thresholds (0–10). Blue lines: MIRACLE2 score; orange lines: Pre-MIRACLE2 model.

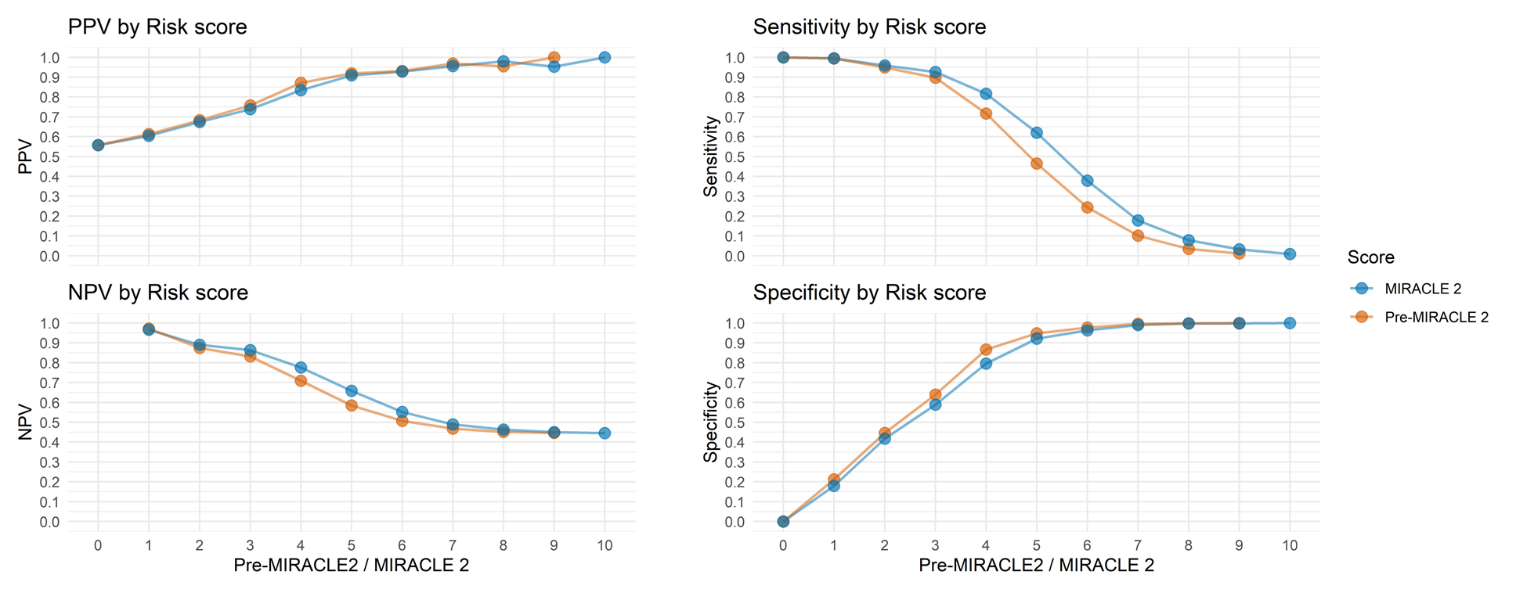

**Supplementary Figure 2. Reclassification of OHCA patients Pre-MIRACLE_2_ to MIRACLE_2_.** This is shown by risk categories (low: 0–2, green; intermediate: 3–4, yellow; high: ≥5, red) and association with cerebral performance category outcome (good: green; poor: red). (Left) Sankey diagram showing patient flows and reclassifications (n, %). (Right) Probability of arterial pH <7.20 by Pre-MIRACLE2 score, with observed proportions.

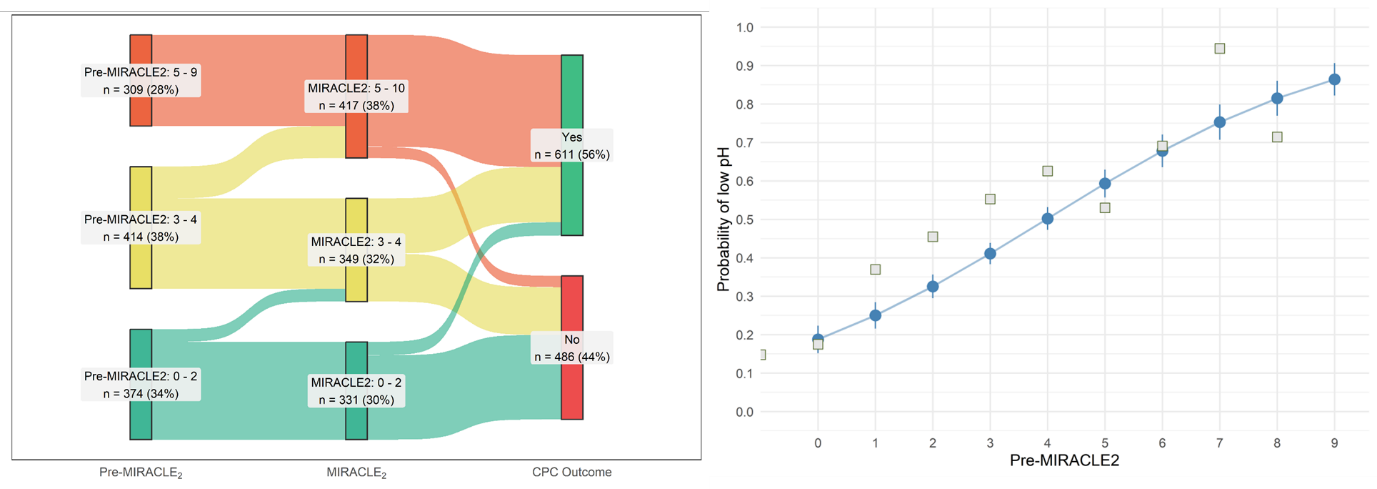
